# Diagnosis in children with exercise-induced respiratory symptoms: a multi-centre study

**DOI:** 10.1101/2020.01.10.20016956

**Authors:** Eva SL Pedersen, Cristina Ardura-Garcia, Carmen CM de Jong, Anja Jochmann, Alexander Moeller, Dominik Mueller-Suter, Nicolas Regamey, Florian Singer, Myrofora Goutaki, Claudia E Kuehni

## Abstract

**Objective:** Exercise-induced respiratory symptoms (EIS) are common in childhood and reflect different diseases that can be difficult to diagnose. In children referred to respiratory outpatient clinics for EIS, we compared the diagnosis proposed by the referring primary care physician with the final diagnosis from the outpatient clinic and described diagnostic tests performed and treatment prescribed after the diagnostic evaluation.

**Design:** Observational study nested in the Swiss Paediatric Airway Cohort (SPAC), which includes respiratory outpatients aged 0-16 years.

**Patients:** We included children with EIS as main reason for referral. Information about diagnostic investigations, final diagnosis, and treatment prescribed came from outpatient records.

**Results:** 214 were referred for EIS (mean age 12 years, 99 (46%) female). The final diagnosis was asthma in 115 (54%), extrathoracic dysfunctional breathing (DB) in 35 (16%), thoracic DB in 22 (10%), asthma plus DB in 23 (11%), insufficient fitness in 10 (5%), chronic cough in 6 (3%), and other diagnoses in 3 (1%). Final diagnosis differed from referral diagnosis in 115 (54%). Spirometry, body plethysmography and measurements of exhaled nitric oxide were performed in almost all; exercise-challenge tests in a third. 91% of the children with a final diagnosis of asthma were prescribed inhaled medication and 50% of children with DB were referred to physiotherapy.

**Conclusions:** Diagnosis given at the outpatient clinic often differed from the diagnosis suspected by the referring physician. Diagnostic evaluation, management and follow-up were inconsistent between clinics and diagnostic groups, highlighting the need for diagnostic guidelines in children seen for EIS.

**Mandatory statements for Archives of Disease in Childhood:** *What is already known on this topic (2-3 statements of max 25 words):*

1. Exercise-induced symptoms are common in childhood but not easy to diagnose because different diagnoses share similar clinical presentations
2. Only few studies focused on children with exercise-induced symptoms and all have included selected groups of patients with difficult-to-diagnose problems

*What this study adds (2-3 statements of max 25 words):*

1. Exercise-induced respiratory symptoms was the main reason for referral in one fifth of the children referred to paediatric respiratory outpatient clinics.
2. Dysfunctional breathing is an under-recognised diagnosis; it was frequently diagnosed in the outpatient clinic (in 37%) but rarely suspected by the referring physician (6%)
3. Diagnostic evaluation, management, and follow-up were inconsistent between clinics highlighting the need for diagnostic guidelines in children seen for EIS.

## Introduction

Exercise-induced respiratory symptoms (EIS) are common in childhood,(1-3) but are not easy to diagnose because different aetiologies share similar clinical presentations.(4-6) EIS are typically due to asthma or exercise-induced bronchoconstriction, but other diseases can cause EIS such as dysfunctional breathing disorders, insufficient fitness level, chronic cough, or rare aetiologies (figure 1).(7, 8) Dysfunctional breathing (DB) disorders are abnormal biomechanical patterns of breathing classified as either extrathoracic (e.g. inducible laryngeal obstruction (ILO)) or thoracic (e.g. pattern disordered breathing).(4, 8) Besides functional causes (e.g. ILO, pattern disordered breathing) dysfunctional breathing can result from structural abnormalities such as laryngomalacia.(9, 10) The diagnosis in children with EIS is complicated by possible coexistence of the different causes (11). When investigating children with EIS a thorough history, physical examination and additional diagnostic procedures are essential. Spirometry and measurement of exhaled nitric oxide are helpful to diagnose asthma, particularly combined with a bronchodilator test.(12) The exercise-challenge test is helpful to reproduce exercise-induced bronchoconstriction or other symptoms reported by the patient and to diagnose ILO.(13) Cardiopulmonary exercise testing monitors gas exchange during exercise and is typically used for proving hyperventilation or an insufficient fitness level, and invasive testing such as flexible laryngoscopy allows to directly visualise laryngeal function during exercise.(1)

**Figure 1:**
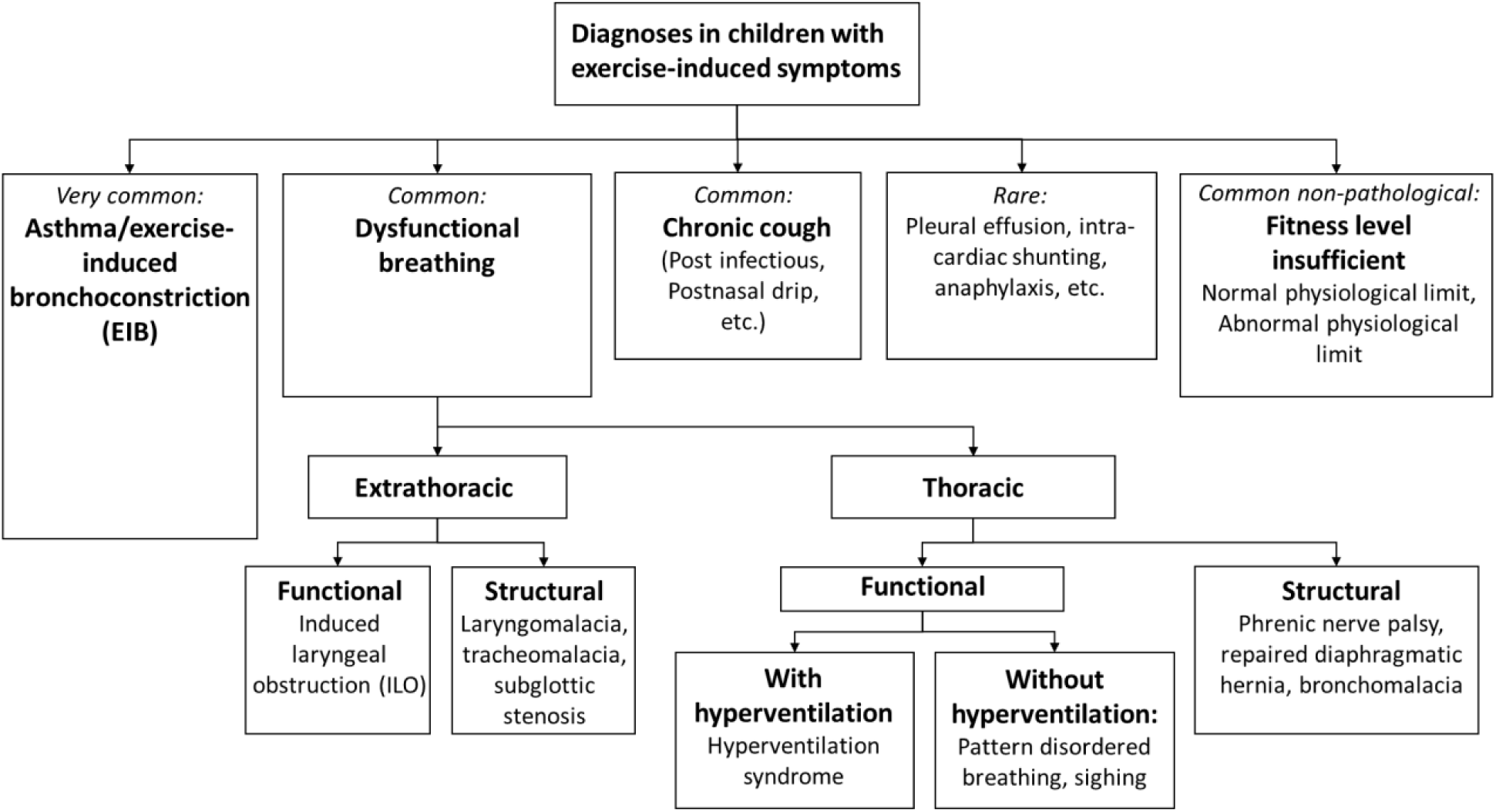
Classification of causes of exercise-induced symptoms used in this study.

Prolonged duration of EIS can lead to physical activity avoidance,(14, 15) reduced quality of life,(16) and overtreatment with inhaled corticosteroids if mistakenly diagnosed as asthma.(6, 17) Only few studies have investigated diagnostic practices and diagnoses given to children seen specifically for EIS(7, 17-21), and all have focused on selected groups of patients excluding children with asthma. No studies have reported the prevalence of different diagnoses and diagnostic practices in representative samples of children with EIS of any cause. We analysed data from Swiss paediatric respiratory outpatient clinics to compare the diagnosis proposed by the referring primary care physician with the diagnosis received at the paediatric respiratory outpatient clinic, and describe diagnostic investigations and treatment prescribed before and at the outpatient clinic.

## Methods

### Study design

We used data from the Swiss Paediatric Airway Cohort (SPAC), an observational national multi-centre clinical cohort from Switzerland.(22) The study included children aged 0-16 years who were referred to the general paediatric respiratory outpatient clinic of participating hospitals for respiratory problems such as wheeze, cough, dyspnoea, sleep- or exercise-related symptoms and spoke sufficient German to participate. Recruitment for SPAC started in July 1, 2017 and is ongoing. By the time we extracted data for this analysis (October 22, 2019), SPAC recruited patients from five paediatric respiratory outpatient clinics in Switzerland. Among 2436 children invited, 1405 (58%) agreed to participate. The SPAC study was approved by the Bern Cantonal Ethics Committee (Kantonale Ethikkomission Bern 2016-02176). Written informed consent was obtained from parents and directly from patients older than 13 years. This paper is reported following the STROBE statement.(23)

### SPAC study procedures and data sources

Eligible patients were recruited at their first clinical visit, where a physician explained the SPAC study. Parents filled in a questionnaire before or shortly after the visit including information on symptoms, medication, environmental exposures and health behaviours. After the visit, the SPAC study team collected referral letters with information on referral diagnosis, and outpatient clinic letters with information on symptoms history, previous treatments, physical examination, diagnostic tests done, and final diagnosis. Results from diagnostic tests were collected from the clinic records and all information was entered into a Research Electronic Data Capture (REDCap) database.(24)

### Inclusion criteria

We included children who were referred for EIS as main referral reason. We considered EIS as main reason for referral if the referral letter or the first outpatient clinic letter described EIS as the only or main reason for referral (supplementary file 1). We excluded children with missing information on referral reason or missing final diagnosis.

### Referral diagnosis

Referral diagnosis was the diagnosis described as suspected cause of EIS in the referral letter from the referring physician. Suspected referral diagnoses were categorised into three categories: asthma (including asthma, recurrent wheeze, or exercise-induced bronchoconstriction); DB (including extrathoracic or thoracic DB); or unknown aetiology if no suspected diagnosis was described.

### Final diagnosis given at outpatient clinic

Final diagnosis was defined as the diagnosis described in the outpatient clinic letter that was sent back to the referring physician after completion of the diagnostic evaluation (which sometimes required more than one visit). Combinations of diagnoses were considered where coexisting diagnoses were listed. We grouped diagnoses into seven categories suggested in previous publications (4, 8) (figure 1). Asthma, extrathoracic DB, thoracic DB, asthma plus any DB, chronic cough, insufficient fitness level, and other diagnoses. We grouped DB into extrathoracic DB (functional: induced laryngeal obstruction, and structural: laryngomalacia, subglottic stenosis) and thoracic DB (functional: pattern disordered breathing, hyperventilation, sighing). For some analyses, we merged rare diagnoses (insufficient fitness level, chronic cough other diagnoses) into one category (supplementary file 1). The final diagnosis was categorised as suspected if the diagnosis in the outpatient clinic letter included the word “suspected”.

### Diagnostic tests performed at outpatient clinic

We extracted information on diagnostic testing from the outpatient clinic letter. Tests included: spirometry, body plethysmography, bronchodilator test, fraction of exhaled nitric oxide (FeNO), allergy tests (skin prick test or specific IgE), chest x-ray, and bronchial challenge tests such as methacholine and exercise-challenge test. Diagnostic tests were performed according to published guidelines (25-27). Challenge tests were often performed at a follow-up visit and we therefore collected challenge tests also from follow-up visits. Children withheld short acting beta2-agonists (SABA) for 8 hours, inhaled corticosteroids (ICS), leukotriene antagonists, and long acting beta2-agonists (LABA) for 24 hours, and antihistamines and sodium cromoglycate for 72 hours before the outpatient clinic visit. All tests were performed by experienced lung function technicians who also assessed quality of the tests.

### Prescribed treatments and other variables

We extracted information about treatment taken prior to the first outpatient clinic visit from the referral letter (described by referring physician) and the first outpatient clinic letter (described in clinical history). Treatment prescribed at the outpatient clinic was taken from the outpatient clinic letter with the latest data and summarised as: SABA, ICS, and LABA or combinations. Information on referral to physiotherapy or other specialty and any planned follow-up visits were taken from the outpatient clinic letter. Information about age, sex, height and weight was taken from the outpatient clinic letter. We calculated body mass index (BMI) as weight (kg) / height*height (cm) and calculated age-adjusted BMI z-scores based on reference values from the World Health Organisation (28), defining overweight as BMI z-score > 1 and obesity as BMI z-score > 2. We used information on parental education, environmental factors and physical activity from the standardised parental questionnaire.

### Statistical analysis

We compared referral diagnosis with final diagnosis, and described asthma treatment prescribed before and at the outpatient clinic. We compared characteristics of children receiving the different diagnoses using chi-square, fisher’s exact and ANOVA tests. Our dataset had few missing values of which the variables parental education (7%) and BMI (2%) had most, and we reported these variables only for children who had valid information. Our main factors of interest (diagnostic evaluations, diagnosis and treatment) had no missing values. We used STATA version 14 for statistical analysis.

## Results

Of the 1065 children who had their first outpatient visit after June 1, 2017, 214 (20%) had EIS as main reason for referral (supplementary file 2). We included data from five clinics. The largest clinic contributed 71 patients and the smallest 26 patients (table 1). On average, children were 12 years old (SD: 3) and 99 (46%) were female (table 2). The commonest referral diagnosis was asthma in 126 (59%); 12 (6%) were suspected to have DB and no diagnosis was proposed in 74 (35%). 89 (43%) had at least one follow-up visit. The average time between baseline and last visit was 3.7 months (range 0.4-16.8).

**Table 1:**
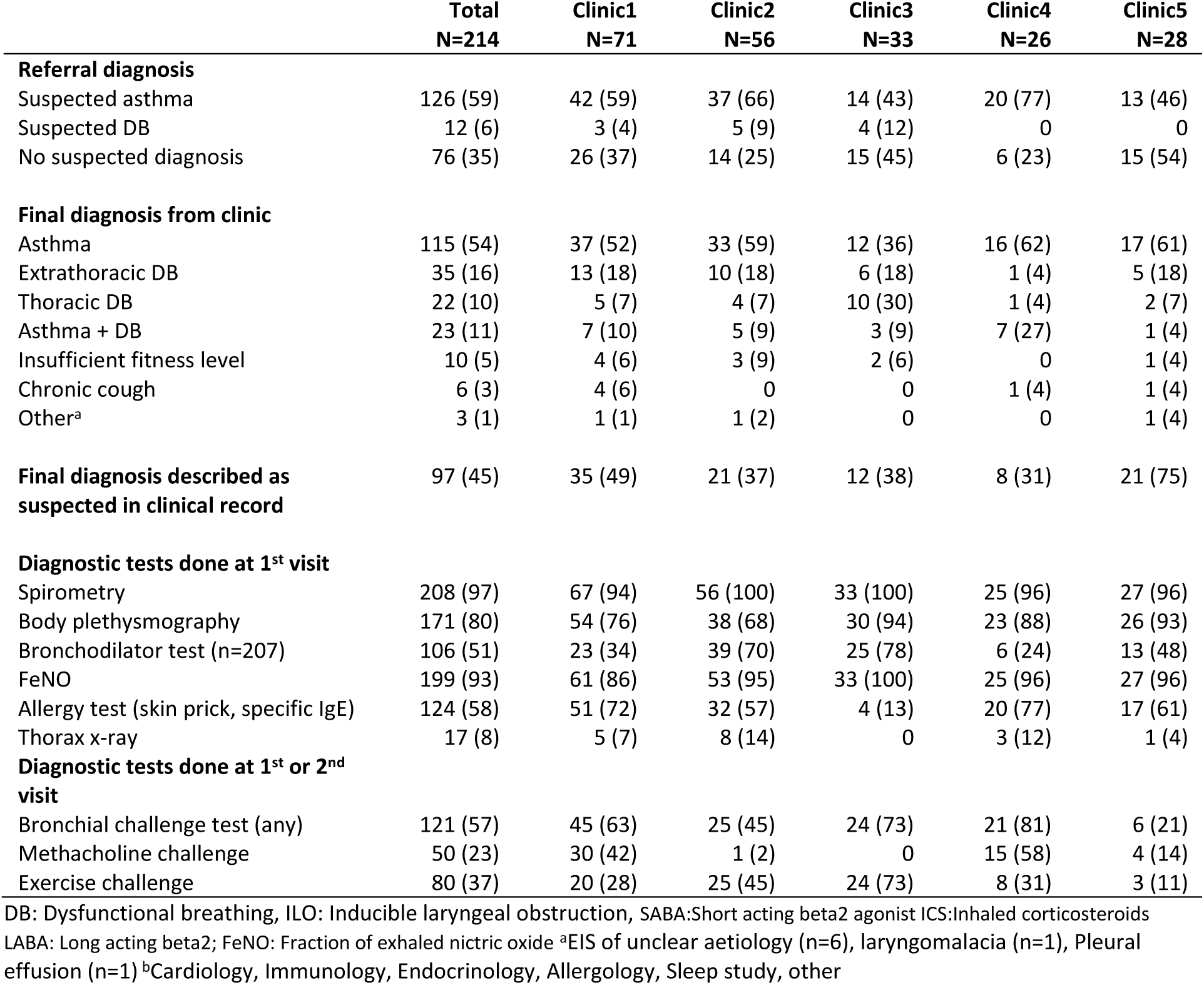
Suspected referral diagnosis, final diagnosis and diagnostic tests described in outpatient clinic letter, in total and by centre (N=214)

|  | **Total**<br>**N=214** | **Clinic1**<br>**N=71** | **Clinic2**<br>**N=56** | **Clinic3**<br>**N=33** | **Clinic4**<br>**N=26** | **Clinic5**<br>**N=28** |
| --- | --- | --- | --- | --- | --- | --- |
| **Referral diagnosis** |  |  |  |  |  |  |
| Suspected asthma | 126 (59) | 42 (59) | 37 (66) | 14 (43) | 20 (77) | 13 (46) |
| Suspected DB | 12 (6) | 3 (4) | 5 (9) | 4 (12) | 0 | 0 |
| No suspected diagnosis | 76 (35) | 26 (37) | 14 (25) | 15 (45) | 6 (23) | 15 (54) |
| **Final diagnosis from clinic** |  |  |  |  |  |  |
| Asthma | 115 (54) | 37 (52) | 33 (59) | 12 (36) | 16 (62) | 17 (61) |
| Extrathoracic DB | 35 (16) | 13 (18) | 10 (18) | 6 (18) | 1 (4) | 5 (18) |
| Thoracic DB | 22 (10) | 5 (7) | 4 (7) | 10 (30) | 1 (4) | 2 (7) |
| Asthma + DB | 23 (11) | 7 (10) | 5 (9) | 3 (9) | 7 (27) | 1 (4) |
| Insufficient fitness level | 10 (5) | 4 (6) | 3 (9) | 2 (6) | 0 | 1 (4) |
| Chronic cough | 6 (3) | 4 (6) | 0 | 0 | 1 (4) | 1 (4) |
| Other<sup>a</sup> | 3 (1) | 1 (1) | 1 (2) | 0 | 0 | 1 (4) |
| **Final diagnosis described as suspected in clinical record** | 97 (45) | 35 (49) | 21 (37) | 12 (38) | 8 (31) | 21 (75) |
| **Diagnostic tests done at 1<sup>st</sup> visit** |  |  |  |  |  |  |
| Spirometry | 208 (97) | 67 (94) | 56 (100) | 33 (100) | 25 (96) | 27 (96) |
| Body plethysmography | 171 (80) | 54 (76) | 38 (68) | 30 (94) | 23 (88) | 26 (93) |
| Bronchodilator test (n=207) | 106 (51) | 23 (34) | 39 (70) | 25 (78) | 6 (24) | 13 (48) |
| FeNO | 199 (93) | 61 (86) | 53 (95) | 33 (100) | 25 (96) | 27 (96) |
| Allergy test (skin prick, specific IgE) | 124 (58) | 51 (72) | 32 (57) | 4 (13) | 20 (77) | 17 (61) |
| Thorax x-ray | 17 (8) | 5 (7) | 8 (14) | 0 | 3 (12) | 1 (4) |
| **Diagnostic tests done at 1<sup>st</sup> or 2<sup>nd</sup> visit** |  |  |  |  |  |  |
| Bronchial challenge test (any) | 121 (57) | 45 (63) | 25 (45) | 24 (73) | 21 (81) | 6 (21) |
| Methacholine challenge | 50 (23) | 30 (42) | 1 (2) | 0 | 15 (58) | 4 (14) |
| Exercise challenge | 80 (37) | 20 (28) | 25 (45) | 24 (73) | 8 (31) | 3 (11) |

DB: Dysfunctional breathing, ILO: Inducible laryngeal obstruction, SABA:Short acting beta2 agonist ICS:Inhaled corticosteroids
LABA: Long acting beta2; FeNO: Fraction of exhaled nitric oxide <sup>a</sup>EIS of unclear aetiology (n=6), laryngomalacia (n=1), Pleural effusion (n=1) <sup>b</sup>Cardiology, Immunology, Endocrinology, Allergology, Sleep study, other

**Table 2:**
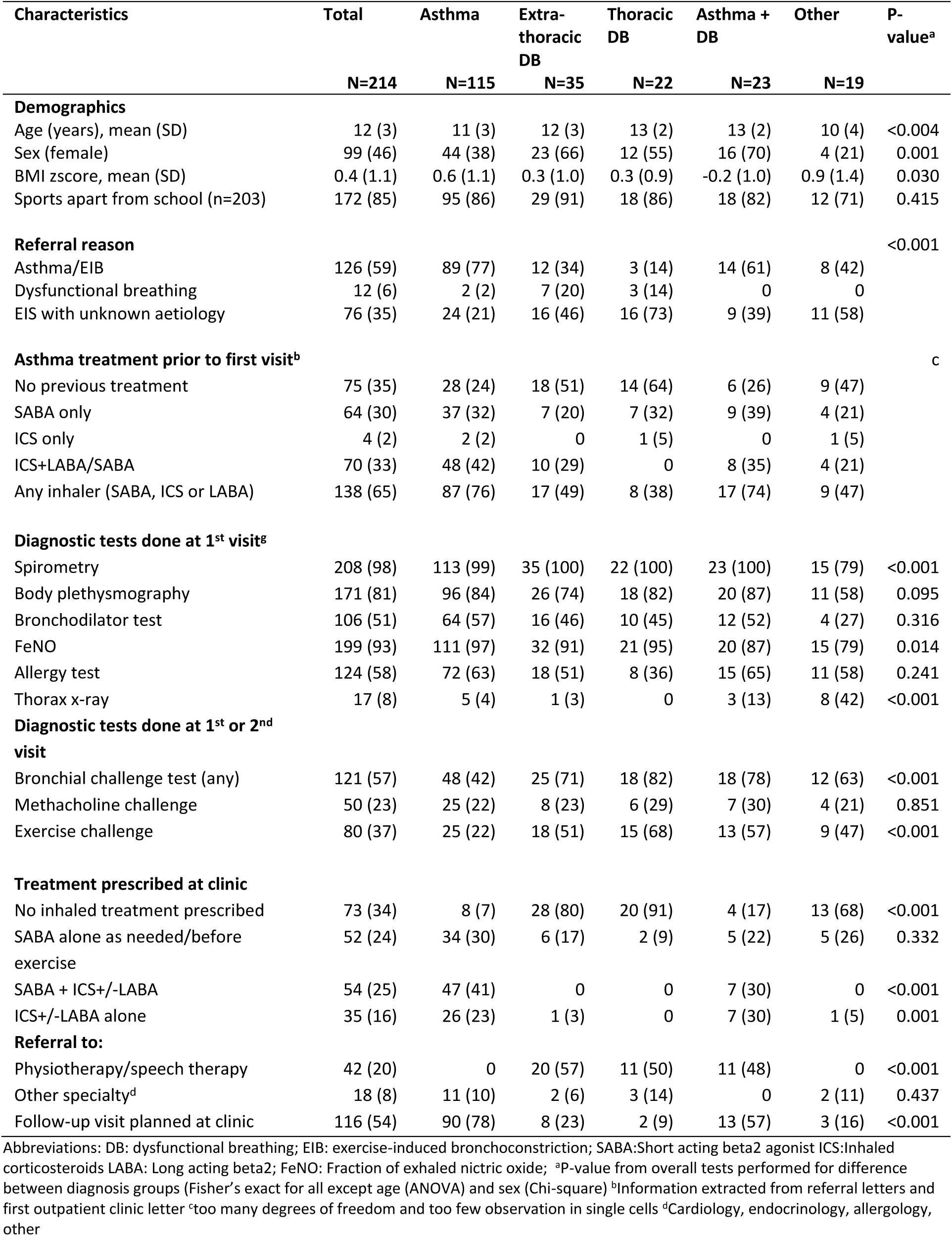
Patient characteristics, referral reason, asthma treatment prior to first visit and diagnostic tests performed at outpatient clinic by final diagnosis.

Final diagnoses from the outpatient clinic letter included asthma (n=115, 54%); extrathoracic DB (n=35, 16%); thoracic DB (n=22, 10%); asthma plus any DB (n=23, 11%), insufficient fitness level (n=10, 5%), chronic cough (n=6, 3%), and other (pleural effusion n=1, unknown aetiology n=2) (table 3). Of the 35 children diagnosed with extrathoracic DB, 32 had functional DB (ILO) and 3 had structural DB. Of the 21 with thoracic DB, all had functional DB (pattern disordered breathing n=16, hyperventilation n=2, sighing tics n=4). In the 23 with asthma plus DB, 19 had asthma plus ILO and 4 had asthma plus pattern disordered breathing. The relative frequency of diagnoses differed between clinics (table 1, supplementary file 3). Children diagnosed with DB or asthma plus DB were slightly older, more often female, and had a lower BMI z-score than children diagnosed exclusively with asthma or other diagnoses. The referral diagnosis often differed from the final diagnosis. Of the 126 referred for suspected asthma, 37 (29%) got another diagnosis at the outpatient clinic (table 2, figure 2). In most (10 of 12) children referred for suspected DB, the diagnosis was confirmed at the outpatient clinic. Of the 76 children with no suspected referral diagnosis, only 24 (32%) were diagnosed with asthma, the majority (n=41) were diagnosed with DB.

**Table 3:** Diagnosis given at outpatient clinic (N=214)

| **Diagnosis** | **n(%)** |
| --- | --- |
| **Asthma** |  |
| Asthma/EIB | 115 (54) |
| **Extrathoracic DB** |  |
| Functional |  |
| ILO | 32 (15) |
| Structural |  |
| Laryngomalacia | 1 (0) |
| Tracheomalacia | 1 (0) |
| Adenoid hyperplasia | 1 (0) |
| **Thoracic DB** |  |
| Functional |  |
| PDB (n=16) | 16 (7) |
| Hyperventilation (n=2) | 2 (1) |
| Sighing tics (n=3) | 4 (2) |
| Structural | 0 |
| **Asthma + DB** |  |
| Asthma+extrathoracic functional DB (ILO) | 19 (9) |
| Asthma+thoracic functional DB (PDB) | 4 (2) |
| **Insufficient fitness level** |  |
| Insufficient fitness level | 10 (5) |
| **Chronic cough** |  |
| Chronic cough unknown aetiology | 4 (2) |
| Post-infectious chronic cough | 2 (1) |
| **Other** |  |
| Bilateral pleural effusion | 1 (0) |
| Unknown aetiology | 2 (1) |

Abbreviations: DB dysfunctional breathing, ILO induced laryngeal obstruction, PDB pattern disordered breathing

**Figure 2:**
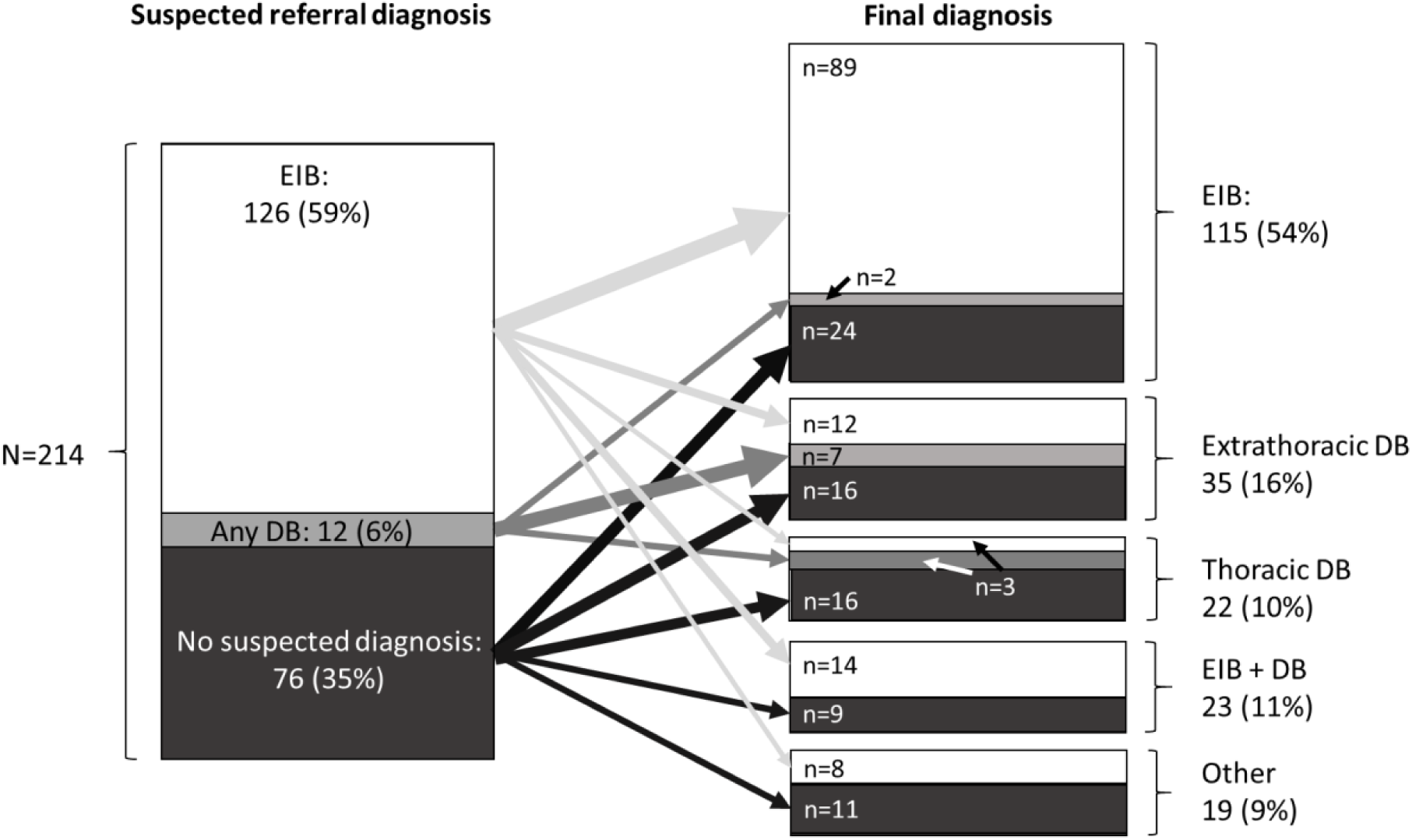
Distribution of suspected referral diagnosis (suspected asthma, suspected dysfunctional breathing (DB), no suspected diagnosis) and final diagnosis (asthma, dysfunctional breathing (DB), asthma + DB, other) with proportions (white, grey, black) indicating relationship between suspected referral diagnosis and final diagnosis.

The diagnostic tests most often performed at the first outpatient clinic visit were spirometry in 208 (97%), body plethysmography in 171 (80%), and FeNO in 199 (93%) (table 1, supplementary file 4). A methacholine challenge test was performed in 50 (23%) and an exercise challenge in 80 (37%). Cardiopulmonary exercise tests or flexible laryngoscopy were not performed. Diagnostic procedures differed by clinic and diagnosis. Children diagnosed with thoracic DB performed exercise-challenge more often (68%) than children diagnosed with EIB (37%) (table 2).

Prior to referral, 65% of all children were on inhaled asthma therapy (30% SABA as needed, 2% ICS and 33% on SABA/ICS or LABA/ICS combinations (table 3). After evaluation at the outpatient clinic, ICS +/-SABA or ICS+LABA was prescribed almost exclusively to children with asthma or asthma plus any DB. SABA alone was mostly prescribed in children with asthma (30%) or asthma plus any DB (22%), but also in those with extrathoracic DB (17%), thoracic DB (9%), and other diagnoses (26%). 42 children (20%) were referred to physiotherapy for breathing/speech training and all of them were diagnosed with extrathoracic or thoracic DB or asthma plus any DB. Follow-up visits were planned in most children (78%) diagnosed with asthma, but only in 23% children diagnosed with extrathoracic DB and 9% with thoracic DB.

## Discussion

This multicentre study of children referred for EIS found that in almost half of the children the diagnosis was revised at the clinic. The commonest final diagnoses apart from asthma were extrathoracic and thoracic DB. Relative frequency of final diagnoses and the set of diagnostic tests performed differed between clinics.

### Strengths and limitations

This pragmatic study is the first to report diagnostic evaluation and management in a real-life clinical setting in children referred to respiratory outpatient clinics for any type of EIS. The broad inclusion criteria (children referred for any type of EIS as main reason for referral) ensured a wide clinical spectrum of children with EIS. Recruitment from five different outpatient clinics in Switzerland made it possible to report on clinical practices and to study variations between different tertiary clinics. A resulting weakness is that diagnostic evaluation and description of final diagnosis were not standardised between clinics, which may influence prevalence estimates. The final diagnosis described in the outpatient clinic letter was described as suspected in 97 (45%), indicating uncertainty in the final diagnosis. In these children, the final diagnosis could change after further diagnostic evaluations, which would influence the prevalence of the estimates.

### Comparison with other studies and interpretation

We identified six previous studies reporting diagnoses given to children seen for exercise-induced symptoms However all six studies included children with EIS unlikely to be caused by asthma (supplementary file 5).(7, 17-19, 21, 29) In our study we included all children with EIS without excluding those with suspected asthma, and for this reason a larger proportion was diagnosed with asthma (57%) compared with previous studies (8-22% asthma). We found that 33 (15%) were diagnosed with ILO, which in previous studies varied between 3-30%. Thoracic DB (e.g. hyperventilation syndrome, sigh dyspnoea, cough), accounted for 10% in our study. In previous studies it varied both in regard to prevalence (4-34%) and labelling of diagnoses, making comparisons difficult. In two previous studies, many patients (19-67%) were diagnosed as having no disease, because their symptoms represented a normal physiological response to exercise with a normal fitness level.(7, 19) In our study, none were diagnosed with normal physiological response to exercise, but ten children were diagnosed with insufficient fitness level. The frequency of diagnoses in our study differed from previous studies, but also differed considerably between clinics (e.g. extrathoracic DB varied from 7% in clinic4 to 47% in clinic3). This suggests a lack of agreement on how to diagnose and define different diagnoses between clinics.

In most children referred for EIS, basic investigations for asthma were performed including measurement of FeNO, allergy tests and lung function testing (spirometry and body plethysmography). Further tests that are diagnostic for other diseases than asthma were done in a minority of children. Exercise challenge testing, recommended to reproduce symptoms in patients with EIS,(4, 26, 30) was only done in 37%. By the time of data collection, none of the clinics performed flexible laryngoscopy and cardiopulmonary exercise test, although laryngoscopy is considered the reference standard for diagnosing extrathoracic DB and cardiopulmonary exercise test is used to diagnose hyperventilation syndrome and insufficient fitness level.(12, 31-33) We found that diagnostic investigations differed between clinics, especially methacholine (0-65%) and exercise challenge tests (7-71%). This indicates little agreement on which diagnostic investigations should be done. Further studies should investigate the optimal algorithm for diagnosing children seen for EIS.

Asthma treatment depends on severity (34) and is therefore expected to differ between children. We would have expected that 100% of the children with asthma would have been prescribed some sort of bronchodilator but in our study, it was only in 93%. Apart from children with asthma, 20% of patients diagnosed with extrathoracic DB were prescribed SABA, which was unexpected but could indicate diagnostic uncertainty. For DB, physiotherapy or speech therapy are recommended treatment.(4, 5) In our study, only half of the children diagnosed with isolated DB (extrathoracic or thoracic) were referred to physiotherapy/speech therapy. The reason for this could be that the pediatric pulmonologist considered the disease as mild and selected a wait-and-see policy after careful instructions about the benign aetiology of the symptoms. Most children diagnosed with asthma (78%) had a planned follow-up visit, but only 23% with extrathoracic DB and 9% with thoracic DB had a planned follow-up visit at the clinic.

In summary, we found that final diagnosis given at the outpatient clinic differed in half of the children from the suspected referral diagnosis. DB was a relatively common diagnosis but rarely suspected by the referring physician. Diagnostic evaluation, management and follow-up were inconsistent between clinics and diagnostic groups. This highlights the need for diagnostic guidelines in children seen for EIS.

## Data Availability

The SPAC dataset is available on reasonable request by contacting Claudia Kuehni by.

## Acknowledgement

We would like to thank the families who took part in the SPAC study. We would also like to thank the outpatient clinic assistants, nurses and doctors for recruiting patients.

## Statement of Ethics

The SPAC study was approved by the Bern Cantonal Ethics Committee (Kantonale Ethikkomission Bern 2016-02176). Written informed consent was obtained from patients’ parents or directly from patients at the age of 14 years and older.

## Disclosure Statement

The authors have no conflicts of interest to declare.

## Funding Sources

This work was funded by the Swiss National Science Foundation (SNSF 32003B_162820) and the Swiss Lung Association. Further funding to develop the SPAC cohort came from the Allergiestiftung U. Müller-Gierok and the Lung league St. Gallen.

## Author’s contributions

EP, CA, CdJ, MG and CK made substantial contributions to the study conception and design. EP drafted the manuscript. EP and CdJ collected and prepared data from the SPAC study. EP, CdJ, CA, AJ, AM, DM, NR, FS, MG, and CK critically revised and approved the manuscript.

## Availability of data and material

The SPAC dataset is available on reasonable request by contacting Claudia Kuehni by.

---

**Supplementary file 1:**
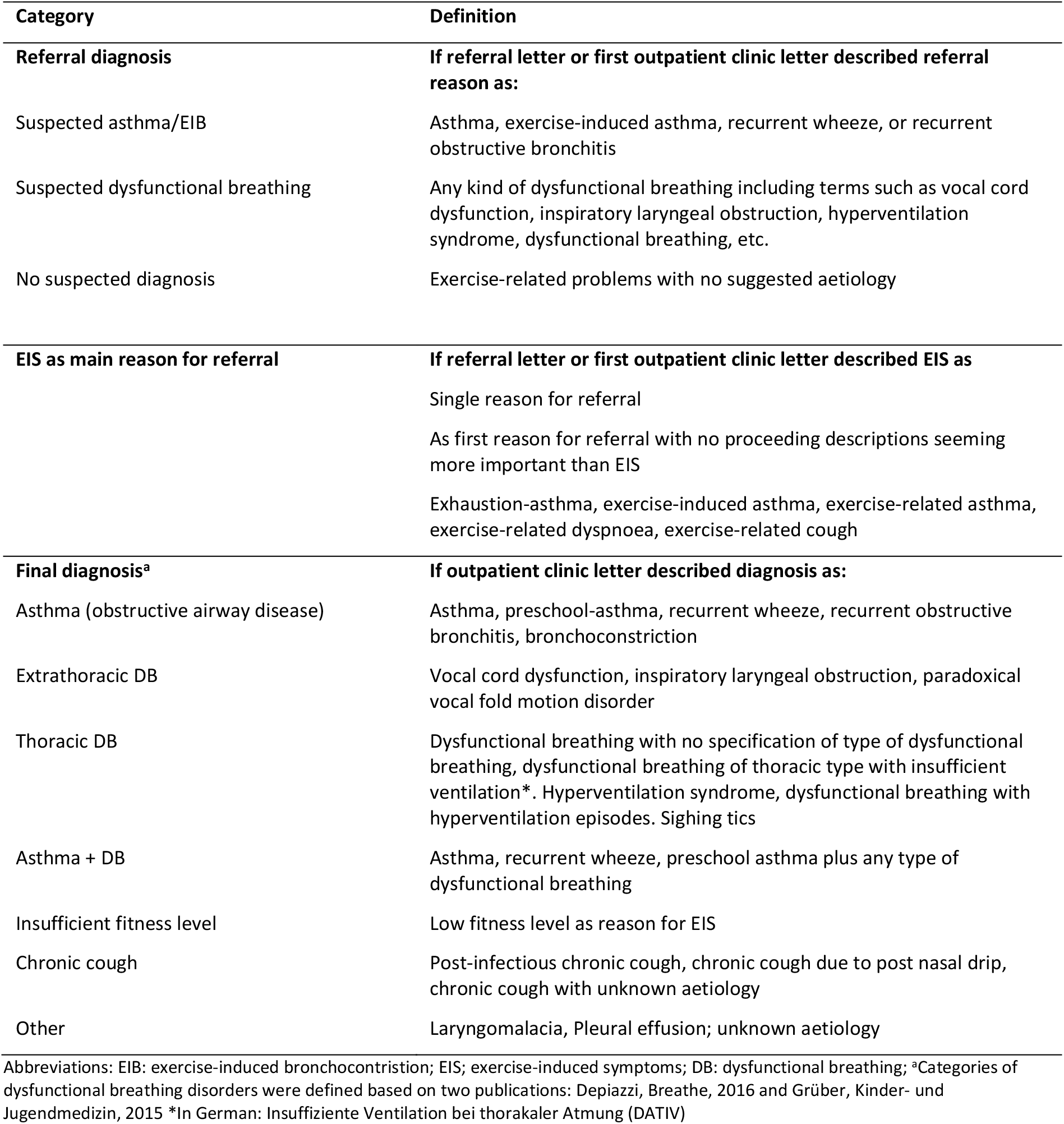
Definitions of variables extracted from referral letters and outpatient clinic letter (terms translated from German)

**Supplementary file 2:**
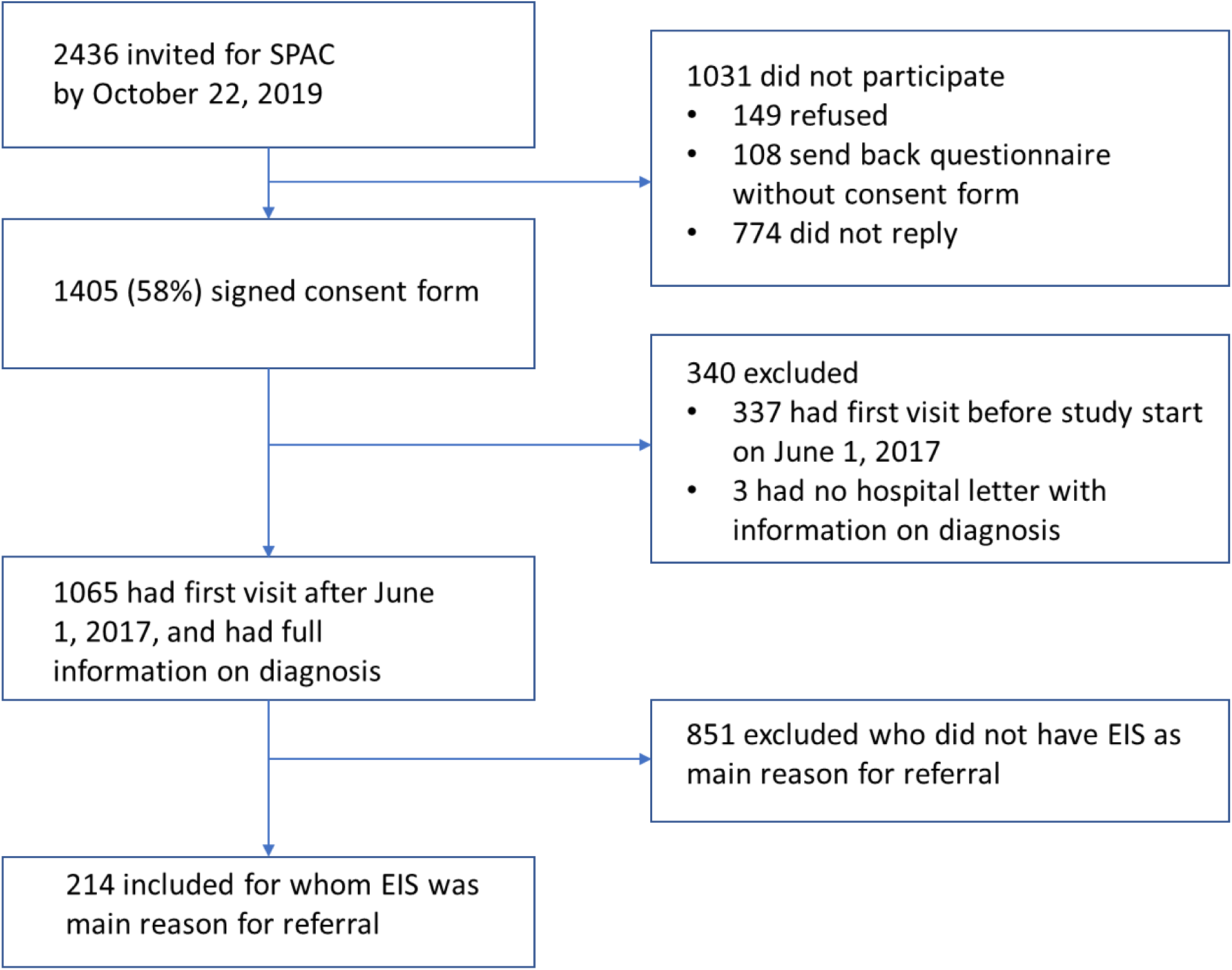
Flow chart of patients included in analysis.

**Supplementary figure 3:**
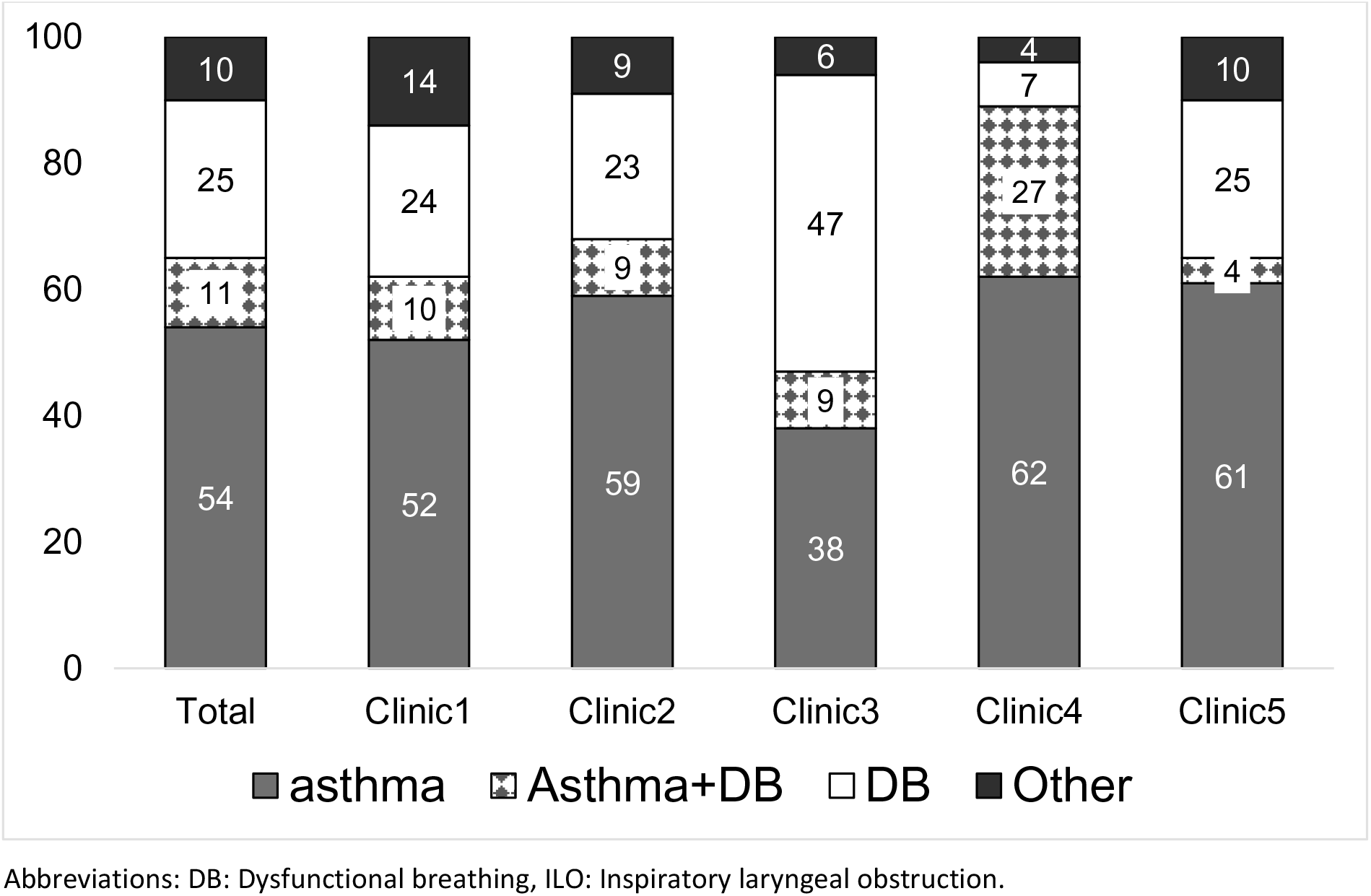
Final diagnosis given to children referred for exercise-induced respiratory symptoms in total population and by clinic. Abbreviations: DB: Dysfunctional breathing, ILO: Inspiratory laryngeal obstruction.

**Supplementary file 4:**
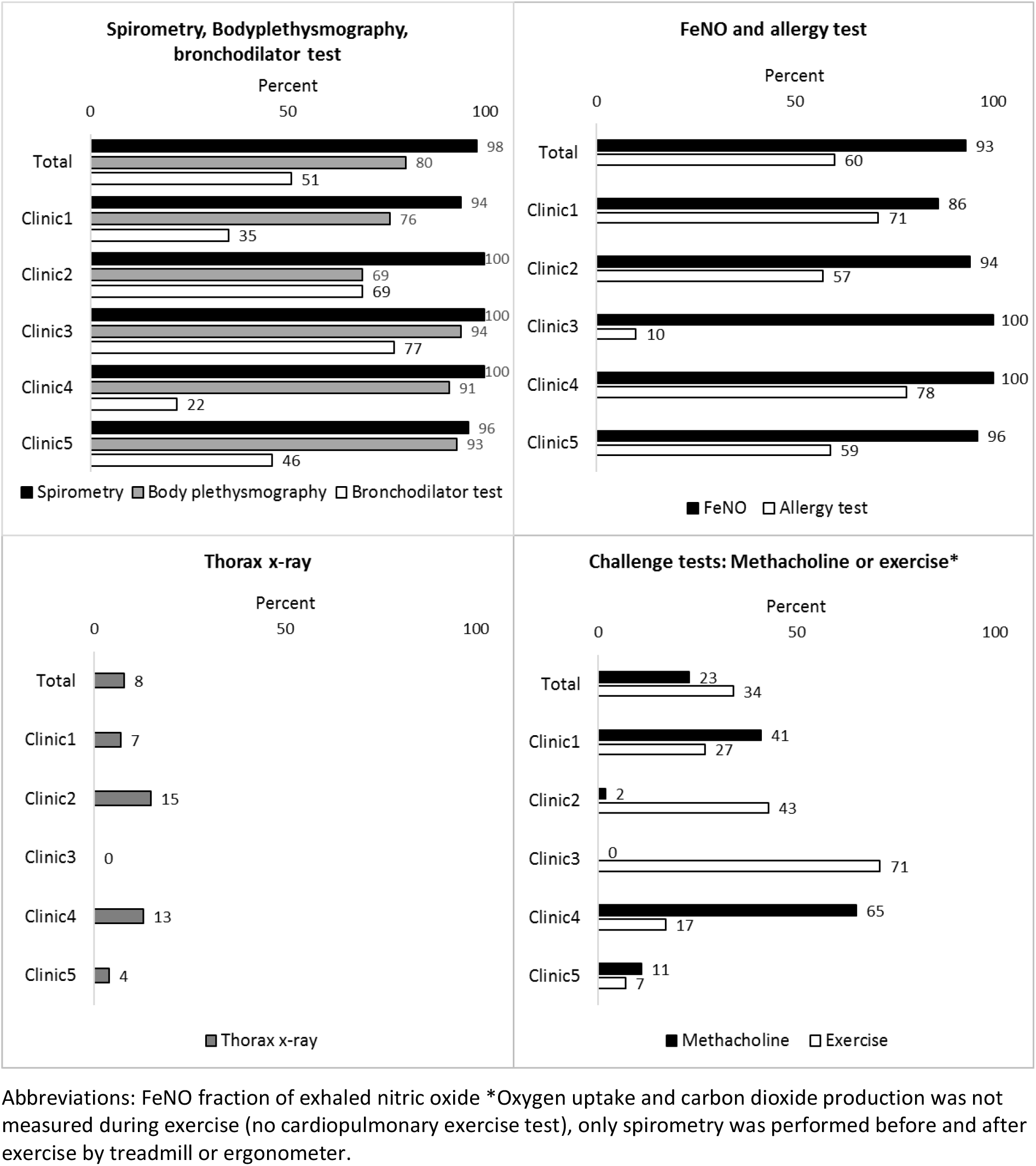
Diagnostic tests performed at outpatient clinic at first visit after referral (spirometry, bodyplethysmography, bronchodilator test, FeNO, allergy test, thorax x-ray) or first or second visit after referral (methacholine- or exercise-challenge test) in total population and by outpatient clinic. Abbreviations: FeNO fraction of exhaled nitric oxide *Oxygen uptake and carbon dioxide production was not measured during exercise (no cardiopulmonary exercise test), only spirometry was performed before and after exercise by treadmill or ergonometer.

**Supplementary file 5:**
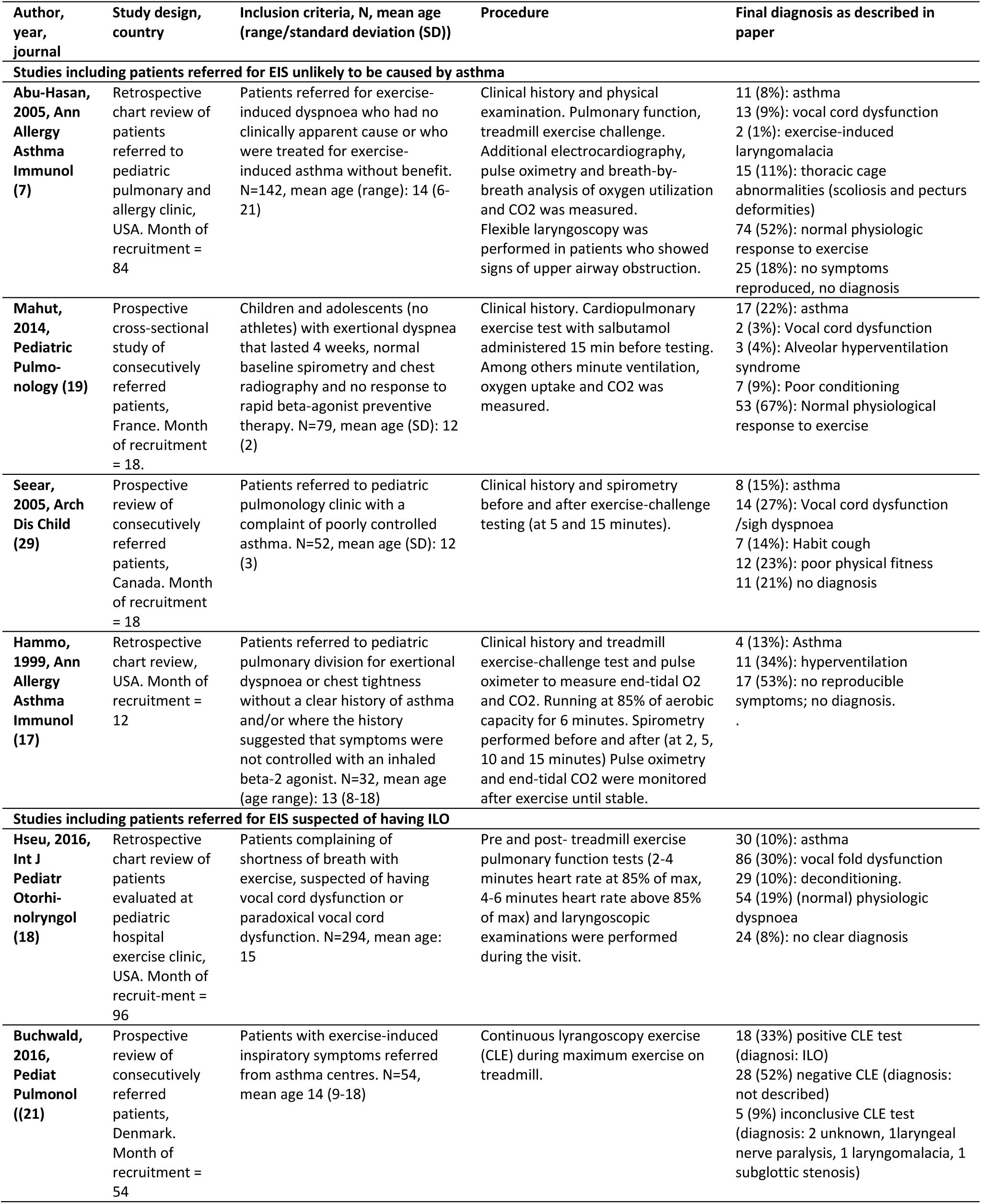
Studies describing diagnosis given to children referred to outpatient clinics for exercise-induced symptoms.

## Notes

### Competing Interest Statement

The authors have declared no competing interest.

